# RT-PCR testing to detect a COVID-19 outbreak in Austria: rapid, accurate and early diagnosis in primary care (The REAP study)

**DOI:** 10.1101/2020.07.13.20152439

**Authors:** Werner Leber, Oliver Lammel, Monika Redlberger-Fritz, Maria Elisabeth Mustafa-Korninger, Karin Stiasny, Reingard Christina Glehr, Eva-Maria Hochstrasser, Christian Hoellinger, Andrea Siebenhofer, Chris Griffiths, Jasmina Panovska-Griffiths

## Abstract

**Background:** Delay in COVID-19 detection has led to a major pandemic. We report rapid early detection of SARS-CoV-2 by reverse transcriptase-polymerase chain reaction (RT-PCR), comparing it to the serostatus of convalescent infection, at an Austrian National Sentinel Surveillance Practice in an isolated ski-resort serving a population of 22,829 people.

**Methods:** Retrospective dataset of all 73 patients presenting with mild to moderate flu-like symptoms to a sentinel practice in the ski-resort of Schladming-Dachstein, Austria, between 24 February and 03 April, 2020. We split the outbreak in two halves, by dividing the period from the first to the last case by two, to characterise the following three cohorts of patients with confirmed infection: people with reactive RT-PCR presenting during the first half (early acute infection) vs. those presenting in the second half (late acute), and people with non-reactive RT-PCR (late convalescent). For each cohort we report the number of cases detected, the accuracy of RT-PCR and the duration of symptoms. We also report multivariate regression of 15 clinical symptoms as covariates, comparing all people with convalescent infection to those with acute infection.

**Findings:** All 73 patients had SARS-CoV-2 RT-PCR testing. 22 patients were diagnosed with COVID-19, comprising: 8 patients presenting early acute, and 7 presenting late acute and 7 late convalescent respectively; 44 patients tested SARS-COV-2 negative, and 7 were excluded. RT-PCR sensitivity was high (100%) among acute presenters, but dropped to 50% in the second half of the outbreak; specificity was 100%. The mean duration of symptoms was 2 days (range 1-4) among early acute presenters, and 4.4 days (1-7) among late acute and 8 days (2-12) among late convalescent presenters respectively. Convalescent infection was only associated with loss of taste (ORs=6.02;p=0.047). Acute infection was associated with loss of taste (OR=571.72;p=0.029), nausea and vomiting (OR=370.11;p=0.018), breathlessness (OR=134.46;p=0.049), and myalgia (OR=121.82;p=0.032); but not loss of smell, fever or cough.

**Interpretation:** RT-PCR rapidly and reliably detects early COVID-19 among people presenting with viral illness and multiple symptoms in primary care, particularly during the early phase of an outbreak. RT-PCR testing in primary care should be prioritised for effective COVID-19 prevention and control.

**Research in Context:** *Evidence before this study:* A comprehensive and effective test-trace-isolate (TTI) strategy is necessary to keep track of current and future COVID-19 infection in the UK and avoid a secondary wave later this year, as society reopens. As part of a wider TTI strategy, it is important to assess the feasibility of COVID-19 testing in primary care. We searched PubMed for implementation of SARS-CoV-2 testing in primary care using the following search terms: (“SARS-CoV-2” OR “COVID-19”) AND “testing” AND (“primary care” OR “general practice”). We did not find any studies that met these criteria.

*Added value of this study:* To our knowledge, our study provides first evidence that extension of a National Influenza Surveillance Programme to include SARS-CoV-2 RT-PCR testing in primary care leads to viral detection among patients presenting with mild to moderate flu-like illness during a local outbreak of COVID-19. We show that the sensitivity of reverse transcriptase-polymerase chain reaction (RT-PCR), the technique to detect viral RNA, is high (100%) in the initial phase of the outbreak and among patients who were acutely unwell. Acute infection was associated with multiply symptoms: loss of taste, nausea and vomiting, breathlessness, myalgia and sore throat; but not loss of smell, fever or cough. We also show high correlation between reactive RT-PCR and seropositivity.

*Implications of all available evidence:* Our findings suggest that RT-PCR can rapidly and reliably detect early COVID-19 among people presenting with viral illness and multiple symptoms in primary care, particularly during the early phase of an outbreak. Furthermore RT-PCR testing in primary care can effectively detect new COVID-19 clusters in the community and should be included in any strategy for prevention and control of the disease.

## Introduction

The COVID-19 pandemic, caused by the novel coronavirus SARS-CoV-2, continues to spread globally with more than 8.5 million cases, and over 450,000 deaths reported as of June 19, 2020. Undetected infection and delays in implementing an effective test-trace-isolate (TTI) strategy have contributed to the spread of the virus becoming a pandemic. SARS-CoV-2 virus has a wide spectrum of manifestations including no symptoms (asymptomatic infection), mild to moderate to severe flu-like illness, pneumonia and acute respiratory distress syndrome (ARDS), sepsis, multi-organ failure and death.^1^ As well as the symptoms associated with flu-like illnesses such as cough, sore throat, fever, fatigue and headaches, altered taste or smell have recently been accepted as markers of COVID-19 infection.^2-4^ In studies to date, the reported time for the infection to become symptomatic (incubation period) varies among different cohorts and settings, with a median incubation period around 5.1 days,^5^ infectivity starting 2.3 days before symptom onset, peaking 1-2 days before that,^6,7^ and gradually declining over 7-10 days.^8,9^

SARS-CoV-2 has the potential for ‘superspreading’ events, resulting in clusters of disease outbreaks among a large number of people.^10^ Although most infections remain isolated cases, a small number of individuals (10%) may cause up to 80% of secondary transmissions.^11^ Undocumented infection may constitute the majority of cases (86%), causing more than half (55%) of all documented infections.^12^ Superspreading events have been reported from across the globe, and countries achieving early viral suppression took rapid and decisive action to implement comprehensive case identification and testing, combined with contact tracing and isolation.^13,14^ For epidemic control of COVID-19, the reproduction number R needs to be less than 1, so that each newly infected person passes the infection on to less than one other person, ensuring infections decline. The presence of undetected and persistent infection within the population, even if very small, can increase R and induce a secondary peak of infections. Therefore, rapid identification and containment of infection is a key factor for the prevention of onward transmission and controlling the virus to protect the public.^15^

In Austria, the first two COVID-19 cases were reported among travelers from Italy from a hotel in the city of Innsbruck on 25 February, 2020. Multiple superspreading events then occurred among tourists visiting Austrian ski resorts, including the town of Ischgl, that are believed to have led to further outbreaks in the tourists’ home countries, including Germany, Denmark and Sweden.^16^ Austria was one of the first countries to adopt comprehensive lockdown measures on March 16, 2020, including protection of vulnerable groups, penalty fees for breaching self-isolation, and the National health hotline 1450 to facilitate testing at acute care settings and *via* mobile units.^17^ The first death from COVID-19 associated complications occurred on 12 March, 2020, and as of July 03, 17,959 cases and 705 COVID-19 related deaths have been reported.

General practice is considered a key partner in case recording, managing high risk groups and delivery of equitable care.^18-21^ The European Centre for Disease Prevent and Control (ECDC) recommended integration of “COVID-19 surveillance with sentinel surveillance of influenza-like illness or acute respiratory infection”.^22^ However, in some countries like the UK and the USA primary care has been largely excluded from the National TTI strategy.^23^ From February 24, 2020, SARS-CoV-2 reverse transcriptase-polymerase chain reaction (RT-PCR) testing was offered to people presenting with mild to moderate flu-like symptoms to any of the 92 sentinel sites (general practices and paediatric practices) participating in the Austrian National Influenza Surveillance Network.^24^ The new service supplemented the existing National health hotline 1450 for people at risk of COVID-19.^25^

The overall aim of this work is to explore whether rapid early RT-PCR testing in primary care can accurately diagnose COVID-19 infection. To attest this we report the outcomes of SARS-CoV-2 RT-PCR testing at a sentinel practice in the ski resort of Schladming-Dachstein, Austria. RT-PCR is an established technique to detect viral RNA from nasopharyngeal sampling used to diagnose COVID-19.^26^ Our study is the first to suggest RT-PCR testing in primary care as an effective method to rapidly, early and accurately diagnose COVID-19 and an important component of an effective TTI strategy. We report the accuracy (via sensitivity and specificity) of rapidly deployed RT-PCR testing in patients presenting with acute infection by comparing it to anti-SARS-CoV-2 antibody status during convalescence in the same geographically defined study cohort. We also report the earliness of viral RNA detection by comparing the duration and number of symptoms among patients presenting during the first half (early presenters) and the second half (late presenters) of the outbreak, measured by the number of days from the first to the last case detected and dividing that period by two. We also identify the key clinical symptoms of acute and convalescent disease and determine a correlation between these.

## Methods

### Setting

This study was set in a sentinel general practice participating in the National Influenza Surveillance Network in the ski resort of Schladming-Dachstein, political subdistrict of Groebming (population 22,829), Austria. The study was conducted during a local COVID-19 outbreak between March to April 2020, where 29 cases detected by RT-PCR were documented. All patients presenting with mild to moderate flu-like illness were included. Following the report of the first cases in Austria, people with flu-like symptoms were advised to call the National health advice hotline 1450 instead of directly presenting to the hospital or general practice. Patients were advised to phone the general practitioner or receive home-testing by mobile testing units, and home self-isolate and self-care.

### Design

We conducted a longitudinal evaluation comprising a pragmatic cohort to examine the impact of SARS-Cov-2 RT-PCR testing on COVID-19 case detection. Between 24 February and 03 April 2020, RT-PCR testing and seropositivity data were collected to compare two groups within this cohort of patients:

- Patients testing RT-PCR reactive at presentation with acute disease
- Patients confirmed anti-SARS-CoV-2 antibody positive during the convalescence phase (confirmed infection).

We define acute disease as the presence of flu-like symptoms combined with reactive SARS-CoV-2 RT-PCR and positive serostatus; and confirmed infection as the presence of convalescent anti-SARS-CoV-2 antibody 3-6 weeks after the acute illness, irrespective of the RT-PCR result.

### Ethics approval

The study used secondary anonymised data for which approval was granted by the University of Graz Research Ethics Committee, Austria (reference number: 32-429 ex 19/20).

### Intervention

Since the winter season 2000/2001, the National Influenza Screening Network has conducted influenza screening for patients attending sentinel general practices and paediatric practices.^24^ Between November and March, participating practices routinely collect nasopharyngeal swabs from patients presenting with flu-like symptoms. Specimens are sent to the Center for Virology, Medical University of Vienna, Austria, for virus isolation on tissue cultures and PCR detection. This surveillance programme allows for near real-time recording of seasonal influenza virus activity in the country. On February 24, 2020, one day before the first two cases were reported, the National Influenza Screening Network was enhanced to include SARS-CoV-2 RT-PCR testing.

Patients with mild to moderate flu-like symptoms calling the study sentinel practice were offered same day appointments for SARS-CoV-2 RT-PCR testing. RT-PCR results were available within 24 hours, and those patients with a reactive outcome were immediately notified by a clinician and advised to self-isolate for a minimum of two weeks following National policy at that time. Repeat follow-up RT-PCR was arranged by the local public health authority (District Captaincy of Liezen, Austria), and people testing non-reactive on repeat RT-PCR were released from self-isolation. After 3-6 weeks, venous blood was obtained to confirm SARS-CoV-2 infection using ELISA IgG and neutralizing antibody assay. We defined the period of the outbreak as the number of days from the first patient to the last patient testing RT-PCR reactive at the practice.

### Outcome measures

We characterise the outbreak using the following testing and clinical outcomes: A) As first outcome we report the diagnostic accuracy (using sensitivity and specificity) of SARS-CoV-2 RT-PCR testing among patients with mild to moderate flu-like symptoms at presentation by comparing it with anti-SARS-CoV-2 antibody during convalescence. We also report any alternative diagnoses for patients testing COVID-19 negative; and hospital admission and death B) As second outcome we report the earliness of RT-PCR testing by comparing the duration and number of symptoms during the first half of the outbreak (early presenters) and during the second half of the outbreak (late presenters) C) As third outcome, we identify the key clinical symptoms associated with RT-PCR reactivity (acute infection) and convalescent seropositivity (confirmed infection) and determine any potential correlation between these stages of disease.

### Clinical data

We obtained anonymous patient data held within the practice computer system. The practice lead clinician (OL) generated a clinical master case report form before extracting pseudonymised patient records into an Excel spreadsheet. EMH and CH verified the accuracy of the data extraction for all patients. Data were stored on a secure server at the Institute of General Practice and Evidence-based Health Services Research, University of Graz, Austria, before sharing it with the study statistician (JPG) using encrypted email and secure storage at the University of Oxford, UK.

### RT-PCR testing

SARS-CoV-2 RT-PCR was performed in scope of the routine surveillance at the Center for Virology, Medical University of Vienna on a Roche LightCycler (http://www.roche.com; Switzerland) using a primer-set provided by TIB MOLBIOL (https://www.tib-molbiol.com/; Germany).^27^

### Enzyme immune linked assays (ELISA)

IgG serostatus assays were performed according to the manufacturers’ protocol using five different commercial test kits of Anti-SARS-CoV-2 IgG enzyme immune linked assays (ELISA) provided by the following companies: EUROIMMUN (EUROIMMUN Medizinische Labordiagnostika AG, www.euroimmun.com),^28^ and EPITOPE DIAGNOSTICS (Immunodiagnostik AG www.euroimmun.com) respectively.^29^ Reagent wells of the Anti-SARS-CoV-2 IgG ELISA are coated with recombinant antigen derived from the spike protein (S1 domain) of SARS-CoV-2. Reagent wells of the EDI™ Novel Coronavirus COVID-19 IgG ELISA are coated with COVID-19 recombinant full length nucleocapsid protein. ABBOTT performed on the Architect platform (ABBOTT LABORATORIES INC., www.abbott.com), DIASORIN (DIASORIN S.p.A, https://www.diasorin.com/home) performed on the LIAISON® platform and ROCHE performed on the cobas e 801 analyzer. The Abbott SARS-CoV-2 IgG assay is a chemiluminescent microparticle immunoassay (CMIA) for the qualitative detection of IgG against a recombinant SARS-CoV-2 nucleoprotein. Results are reported in form of an index value (S/C). LIAISON® SARS-CoV-2 S1/S2 IgG assay is a chemiluminescence immunoassay (CLIA) for the quantitative detection of IgG against the recombinant S1 and S2 domain of the spike protein. Results are reported in arbitrary units (AU/mL). Elecsys® Anti-SARS-CoV-2 assay (Roche Diagnostics) is a electrochemiluminescence immunoassay (ECLIA) for qualitative detection of CoV2 antibodies in human serum against a recombinant nucleocapsid protein of SARS-CoV-2. It is a total antibody assay not differentiating between IgA, IgM or IgG but detecting IgG predominantly. Results are reported as numeric values in form of signal sample /cutoff (COI).

### Neutralising antibody assay

Samples with discordant antibody results (see below) were further evaluated using an in-house neutralising antibody assay as follows: Serial dilutions of heat-inactivated serum samples were incubated with 50-100 TCID50 SARS-CoV-2 (GISAID/EPI_ISL_438123/hCoV-19/Austria/CeMM0360/2020) for 1h at 37 °C. The mixture was added to Vero E6 (ATCC ® CRL-1586) cell monolayers and incubation was continued for two to three days. NT titers were expressed as the reciprocal of the serum dilution that protected against virus-induced cytopathic effects. NT titers ≥10 were considered positive. The study has been reported in accordance with STARI reporting guidelines for implementation studies.^30^

### Statistical analysis

We present a descriptive statistics of patient demographics including age, gender and ethnicity; and the following three outcomes:

#### Outcome 1

We explored the diagnostic accuracy of the RT-PCR, by determining its sensitivity and specificity. To do this, we stratified RT-PCR results in four groups: true reactive (RT-PCR reactive and confirmed antibody positive); false reactive (RT-PCR reactive and antibody negative); true non-reactive (RT-PCR non-reactive, antibody negative); and false non-reactive (RT-PCR non-reactive, antibody positive).

#### Outcome 2

We calculated the earliness of RT-PCR testing by projecting the mean duration of symptoms, in days (range), and mean number of symptoms (range), across the three cohorts of patients with confirmed infection: early acute, late acute and late convalescent. The three cohorts were obtained by stratifying people with confirmed infection according to the date of presentation to the practice during the outbreak as follows: people presenting with acute infection (RT-PCR reactive, confirmed antibody positive) during the first half of the outbreak (early acute disease) *vs*. those people presenting during the second half of the outbreak (late acute); and those people presenting with convalescent disease (RT-PCR non-reactive but confirmed antibody positive) in the second half of the outbreak (late convalescent).

#### Outcome 3

Multivariate logistic regression explored the association of 15 clinical symptoms with testing RT-PCR reactive at presentation and among all patients with confirmed infection. We reported the odds ratios (ORs) and the significance value (*p*) of each covariate on testing positive in either case. We quantified the association between patients with reactive RT-PCR (and confirmed antibody positive) and all patients with confirmed infection, projecting the correlation coefficient *r*, and the 95% CI.

## Results

### Overall testing results

Baseline characteristics for both subgroups were similar for sex, age, and ethnic origin. Figure 1 shows the flow-chart for the patient cohorts of this study. 73 patients presented with mild to moderate flu-like illness, all of whom received SARS-CoV-2 RT-PCR (and influenza PCR) testing. Of those, 16 (21.9%) tested RT-PCR reactive *vs*. 57 (78.1%) who tested non-reactive including four that tested influenza PCR reactive. Due to lack of venous blood sampling, antibody data was not available for 7 patients (1 RT-PCR reactive vs. 6 non-reactive) that were excluded from this analysis. Therefore, of the 66 patients included in this analysis, 22 patients (33.3%) had Covid-19 infection confirmed by antibody testing and 44 (66.7%) patients were confirmed seronegative. Of all 22 patients with confirmed infection, eight (early acute presenters) presented in the first half of the outbreak (12 days from March 11 to 22, 2020) and 14 patients presented in the second half (March 23 to April 03, 2020); of the latter, seven patients were late acute and seven late convalescent. Alternative diagnoses of the 44 patients who tested SARS-CoV-2 negative included: Influenza and infectious mononucleosis (N=2, each); bacterial tonsillitis, bacterial pneumonia, bronchitis and exacerbation of chronic obstructive pulmonary disease (COPD) (N=1, each) (see flow-chart, Figure 1). No hospital admissions or deaths were reported.

**Figure 1:**
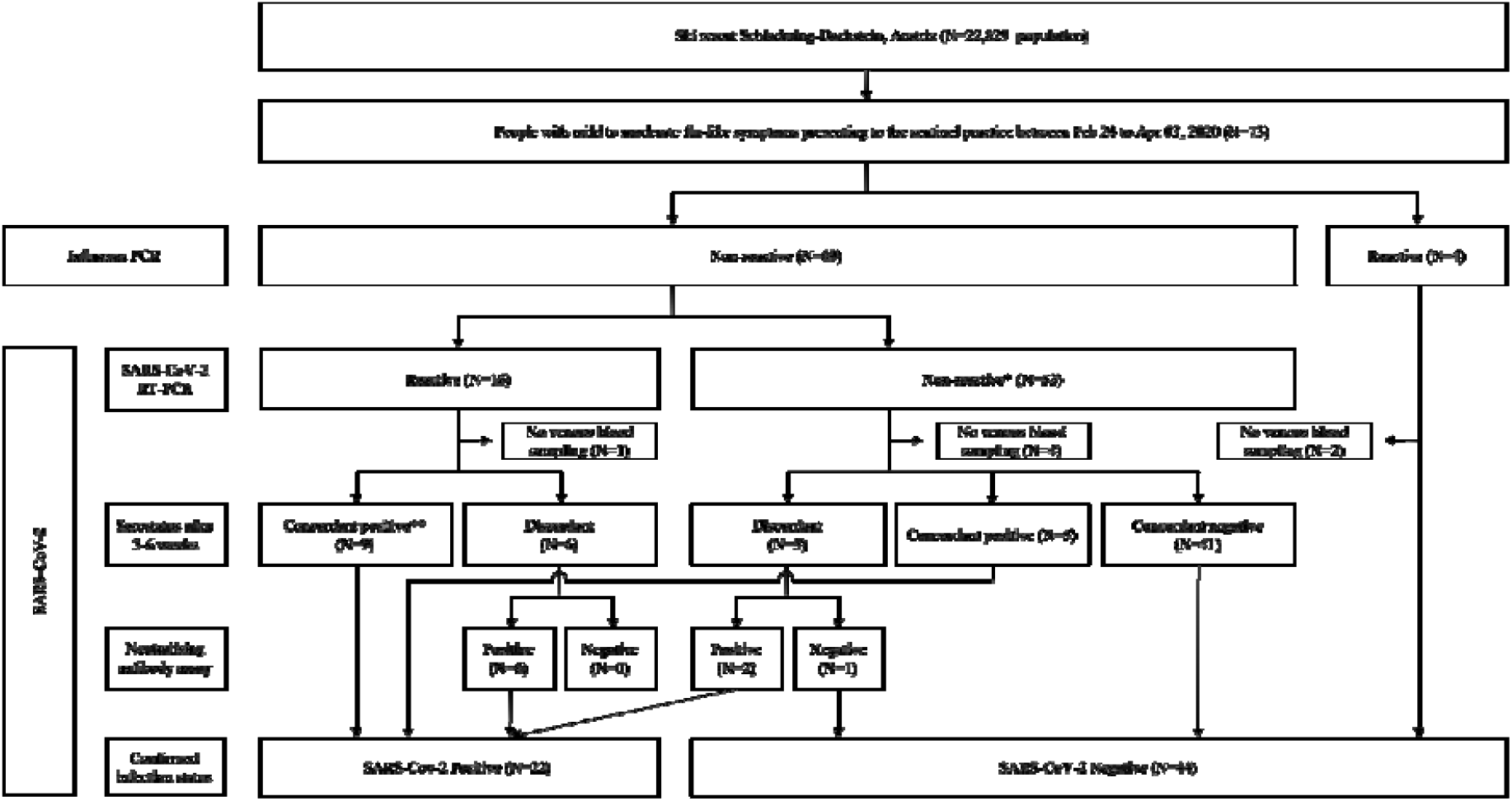
Flow-chart. 22 patients had COVID-19 infection confirmed by antibody testing, including 15 patients diagnosed with acute disease (reactive RT-PCR) and 7 with convalescent disease (non-reactive RT-PCR); among the former, 9 patients tested concordant antibody positive and 6 patients tested neutralizing antibody positive following discordant ELISA result; and among the latter, 5 patients tested concordant antibody positive and 2 patients tested neutralizing antibody positive following discordant ELISA result. 44 patients with non-reactive RT-PCR tested antibody negative, including 41 with concordant negative ELISA, 1 patient with negative neutralizing antibody after discordant ELISA result and 2 patients diagnosed with Influenza. Antibody status was not available for 7 patients. **Final clinical diagnoses included infectious mononucleosis (N=2); bacterial tonsillitis, bacterial pneumonia, and bronchitis and exacerbation of COPD (N=1, each). ***No concordant negatives.

### Specificity and sensitivity of RT-PCR

In the absence of a gold standard, we used a consensus statement on serostatus, irrespective of RT-PCR outcomes, to establish whether an infection had occurred. We considered an infection as confirmed in any patient who tested IgG ELISA positive on all five screening platforms (concordant results) or in any patient with mismatch between ELISA test results (discordant results) but positive neutralising antibody assay (see flow-chart, Figure 1). Of the 15 patients with reactive RT-PCR, sera from nine patients were concordant positive and six were discordant; and of the 53 patients with non-reactive RT-PCR, sera from 41 patients were concordant negative, 5 were concordant positive, and three were discordant. Sera from two patients diagnosed with Influenza, who tested RT-PCR non-reactive, were concordant negative and included in this analysis. For the nine patients with discordant results, we used neutralising antibody assay to confirm infection status. All patients (N=6) with reactive RT-PCR were neutralising antibody positive; and of the 3 patients with non-reactive RT-PCR, two were neutralising antibody positive, and one was negative. Therefore, overall, when combining ELISA and neutralising antibody assay, 22 patients had confirmed infection, of whom 15 patients were RT-PCR reactive (true reactive) and 7 were non-reactive (false non-reactive). There were no false reactive RT-PCR results. Therefore, RT-PCR correctly identified infection in 15/22 patients (overall sensitivity of 68.1%). RT-PCR among all acute (early and late) presenters and during the first half of the outbreak was high (100%), but dropped to 50% in the second half of the outbreak. RT-PCR correctly identified absence of infection for all 44 patients testing antibody negative (true non-reactive) indicating specificity of 100%.

### Earliness of RT-PCR testing

The mean duration of symptoms was 2 days (range 1-4) among early acute presenters, 4.4 days (range 1-7) among late acute presenters, and 8 days (range 2-12) among people with late convalescent infection; and 3.9 days (range 1-14) among non-Covid-19 controls (Figure 2C). The mean number of symptoms was 6.75 (range 4-9) among early acute presenters, 6.86 (3-12) among late acute presenters and 6.3 (1-11) among people with convalescent infection; and 5.23 (range 2-11) among non-Covid-19 controls (Figure 2C).

**Figure 2A:**
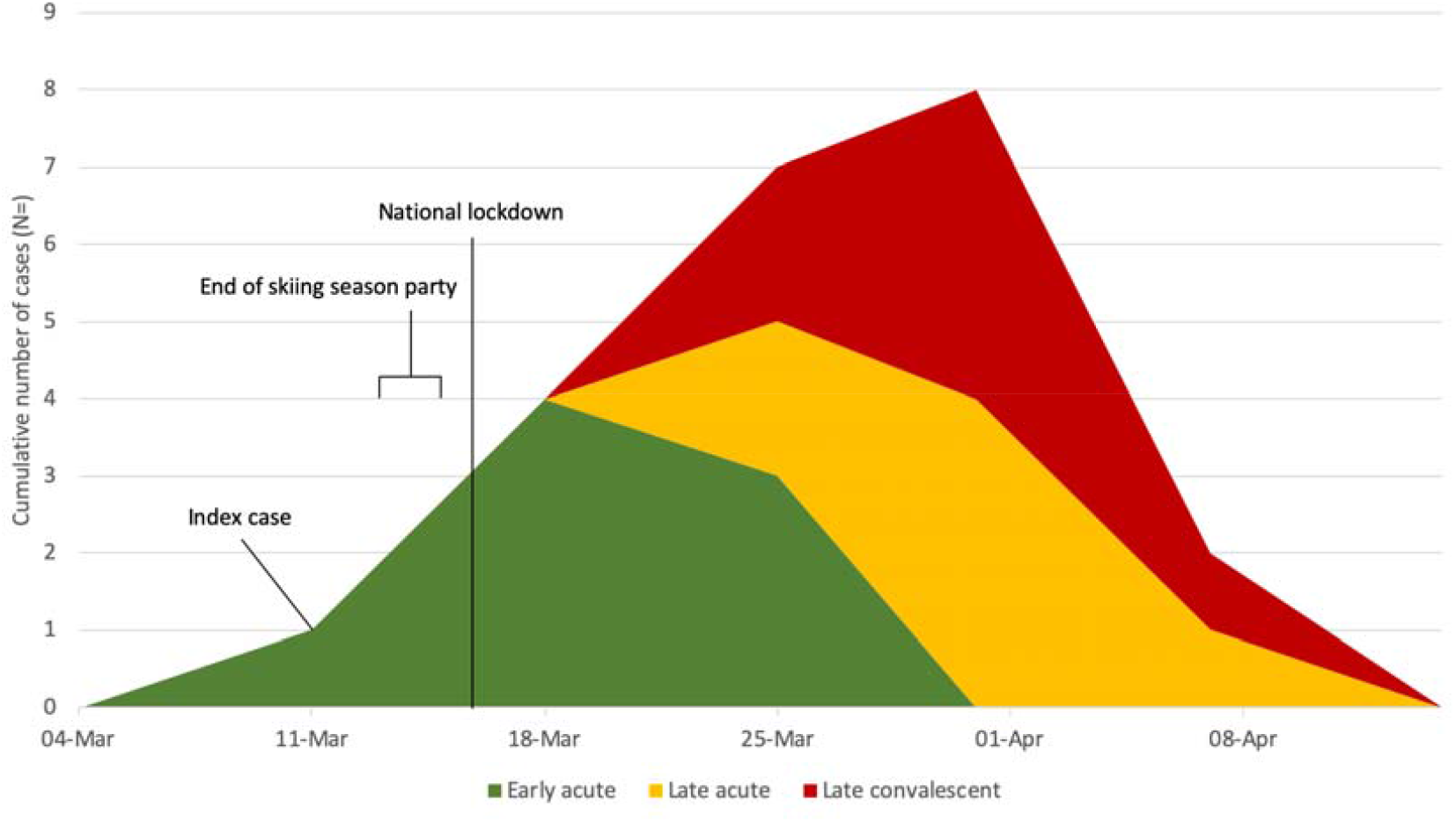
Cumulative COVID-19 diagnosis in the ski-resort Schladming-Dachstein over time. The main outbreak occurred after a three-day event (March 13 to 15) celebrating the early termination of the skiing season due to National lockdown commencing on March 16. Between March 11 (index case) and April 03, 8 people were diagnosed with acute infection (RT-PCR-reactive, confirmed antibody positive) in the first half (12 days from March 11 to 22, 2020) of the outbreak (green colour), and 7 people with late acute infection (amber) and 7 people with convalescent infection (red) were detected during the second half.

**Figure 2B:**
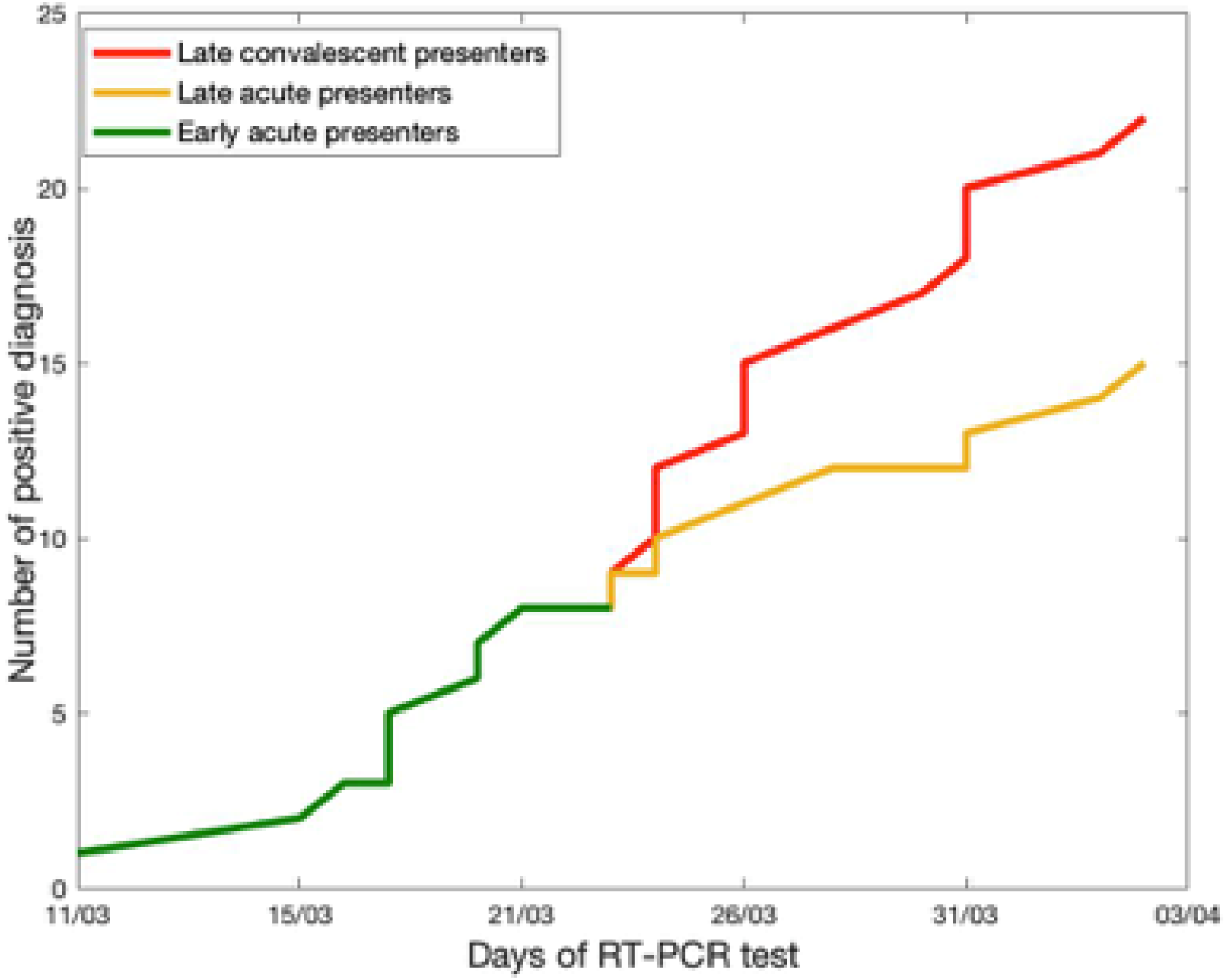
Cumulative weekly numbers of confirmed COVID-19 cases during the outbreak. Timing of patient presentation during the outbreak (N=22) split into people presenting with early acute disease during the first half (12 days) of the outbreak (green colour, N=8), and those presenting with late acute (amber, N=7) and late convalescent disease (red, N=7) in the second half of the outbreak. RT-PCR was 100% sensitive among all early acute and late acute presenters. RT-PCR did not detect any of the late convalescent presenters.

**Figure 2C:**
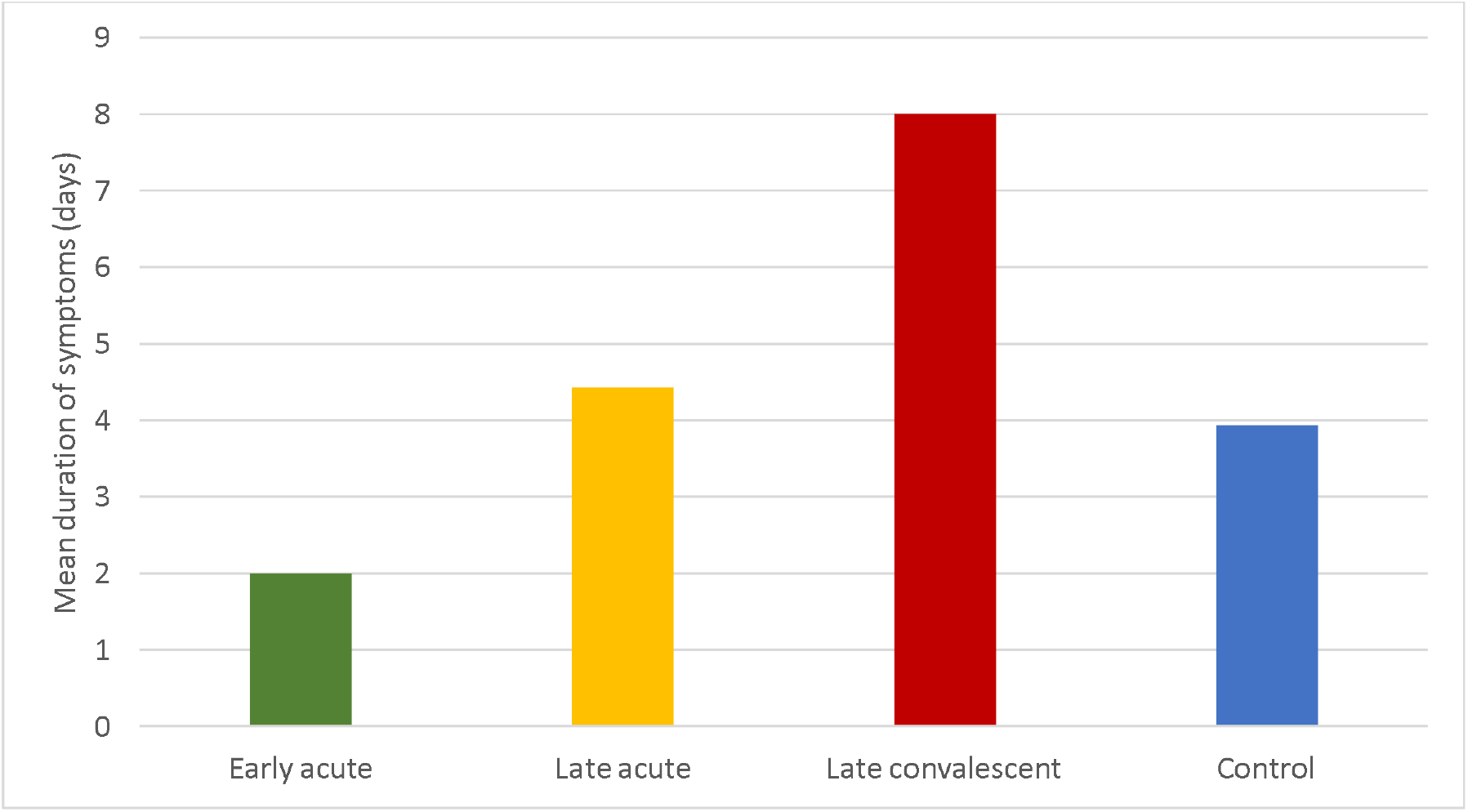
Mean duration of symptoms across people with confirmed infection (N=22) split into those people presenting with early acute infection (green colour, N=), and late acute (amber, N=7) and late convalescent infection (red, N=7); and non-Covid-19 controls (blue, N=44).

**Figure 2D:**
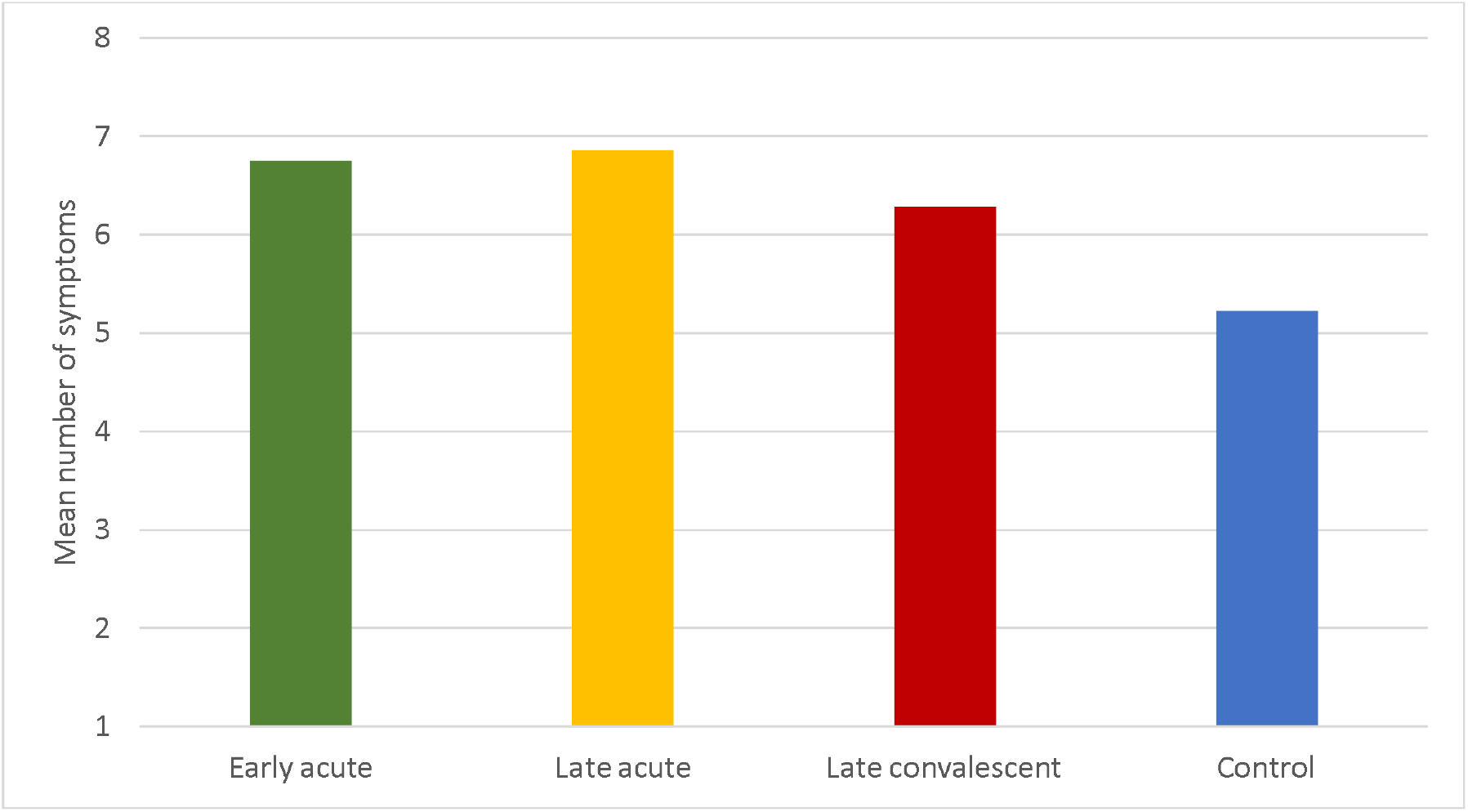
Mean number of symptoms across people with confirmed infection (N=22) split into those people presenting with early acute infection (green colour, N=8), and late acute (amber, N=7) and late convalescent infection (red, N=7); and non-Covid-19 controls (blue, N=44).

### Regression analysis on confirmed infection

Multivariate regression on all 66 patients, including 22 (31.9%) with confirmed infection, suggested that loss of taste, but not loss of smell, was the key covariate significantly associated with positive serostatus (ORs=6.03; p=0.047). (Table 1) Breathlessness (OR=6.9, p=0.054) and cough (OR=0.12, p=0.053) were also possible covariates of confirmed infection.

**Table 1:** Results from the regression analysis of 15 patients presenting with acute SARS-CoV2 infection (RT-PCR reactive, antibody confirmed positive); and 22 patients with confirmed infection (antibody confirmed positive, irrespective of RT-PCR result).

| People with confirmed infection (seropositive irrespective of RT-PCR result) (N=22) |  |  |  |
| --- | --- | --- | --- |
| Clinical symptom | Odds ratio | 95% CI | p-value |
| Change in taste | 6.02 | (1.02,35.51) | 0.047 |
| Nausea and vomiting | 4.42 | (0.748,26.09) | 0.101 |
| Sore throat | 0.36 | (0.067,1.93) | 0.233 |
| Myalgia | 1.15 | (0.24,5.51) | 0.865 |
| Breathlessness | 6.90 | (0.96,49.40) | 0.054 |
| Change in smell | 0.77 | (0.098,6.15) | 0.811 |
| Fever | 2.97 | (0.44,20.35) | 0.266 |
| Cough | 0.12 | (0.014,1.03) | 0.053 |
| People with acute disease (RT-PCR reactive and seropositive) (N=15) |  |  |  |
| Clinical symptom | Odds ratio | 95% CI | p-value |
| Change in taste | 571.72 | (1.92,170629.2) | 0.029 |
| Nausea and vomiting | 370.11 | (2.71,50429.42) | 0.018 |
| Sore throat | 0.002 | (0.000006,0.74) | 0.039 |
| Myalgia | 121.82 | (1.52,9749.08) | 0.032 |
| Breathlessness | 134.46 | (1.02,17796.87) | 0.049 |
| Change in smell | 0.37 | (0.008,15.87) | 0.607 |
| Fever | 1.44 | (0.057,36.66) | 0.825 |
| Cough | 0.011 | (0.00008,1.42) | 0.069 |

### Regression analysis on acute disease

All 15 patients with acute disease reported fatigue and therefore, this covariate was removed from the analysis; and observations from two patients with non-reactive RT-PCR, who did not report fatigue, were also removed (Table 1). The multivariate logistic regression on the remaining 66 patients showed that the following covariates were associated with acute disease: loss of taste (OR=571.72; p=0.029), nausea and vomiting (OR=370.11; p=0.018), breathlessness (OR=134.46; p=0.049), myalgia (OR=121.82; p=0.032) and sore throat (OR=0.002, p=0.039); and but not loss of smell (OR=0.37, p=0.607), fever (OR=1.44, p=0.825) or cough (OR=0.01, p=0.069).

### Correlation between acute and confirmed infection

Testing RT-PCR reactive was correlated with testing seropositive for COVID-19 infection (r=0.77 (95%CI=(0.65,0.89)). Among early and acute presenters, the correlation between the two tests was perfect (green and amber in Figure 2(a)), irrespective of the stage of the outbreak; whereas in the second half of the outbreak, RT-PCR did not detect any case with convalescent infection (red curve on Figure 2(a)).

## Discussion

Our results demonstrate that SARS-CoV-2 RT-PCR testing, when added to a National influenza surveillance programme in primary care, can rapidly, early and accurately diagnose COVID-19 infection. In a cohort of 73 patients tested in the course of the outbreak, 22 patients were diagnosed with COVID-19, comprising: 8 patients presenting early acute, and 7 presenting late acute and 7 late convalescent respectively; 44 patients tested SARS-COV-2 negative, and 7 were excluded. Overall, RT-PCR was 100% specific; and sensitivity among all acute presenters was 100%, but dropped to 50% in the second half of the outbreak. Testing RT-PCR reactive showed perfect correlation with seropositivity during the first half of the outbreak and among early acute and late acute presenters. RT-PCR did, however, not detect any late convalescent presenters that were identified by antibody testing in the second half of the outbreak. Strikingly, the mean duration of symptoms of early presenters (2 days) was less than half of late acute presenters (4.4 days) and a quarter of late convalescent presenters (8 days). These findings highlight the need to undertake RT-PCR testing rapidly and early as soon as symptoms occur. The mean number of symptoms was higher among people with COVID-19 compared to controls. While loss of taste, nausea and vomiting, breathlessness, myalgia and sore throat were strongly associated with acute infection; loss of smell, fever and cough were not. Surprisingly, loss of taste, but not any other clinical symptom, was significantly associated with convalescent infection.

Our study highlights the importance of early RT-PCR testing in primary care among patients presenting with multiple flu-like symptoms. Our results agree with the findings of the King’s College group that loss of taste is an overall marker of COVID-19 infection.^31^ However, our study suggests that the presence of multiple symptoms among patients presenting shortly after symptom onset might indicate acute COVID-19. Furthermore, unlike the Kings’ College study, where the suggestion is made that people use a mobile phone application to self-diagnose and self-isolate, we suggest that people with acute COVID-19 can be accurately diagnosed by RT-PCR in primary care. Given the high accuracy of RT-PCR during acute SARS-CoV-2 infection, a paradigm shift in the management of flu-like illness in primary car may be needed: active testing of patients presenting within the first 1-2 days of showing multiple symptoms should be encouraged rather than self-isolation of symptomatic people. This is aligned to the suggestions in a recent Lancet Editorial.

The outbreak we explored occurred after a three-day party (March 13-15) to celebrate the premature termination of the skiing season due to the Austrian National lockdown measures on March 16. The index case was diagnosed on March 11 and the first secondary cases were reported two days after the celebrations. No other endemic cases were documented in the region following the last case reported in this study. Superspreading events have been associated with high intensity aerosol producing activities (shouting, singing) in confined spaces and paradoxically, the lockdown celebrations might have triggered the local outbreak. Considering the short incubation period of the virus and high infectiousness around onset of disease,^32^ the bulk of cases observed during the first half of the outbreak likely suggests an initial wave of infections among acutely unwell people following high density aerosol exposure at the party. Cases identified in the second half of the outbreak might represent a mix of secondary acute infections, likely occurred in people’s homes, given the strict lockdown policies, and late presentation of people with convalescent disease. Diversity of cases in the second half might explain the drop in sensitivity of RT-PCR from 100% initially by a half later in the outbreak. Hence, our data suggest that nasopharyngeal sampling should be conducted immediately after symptom onset to increase the likelihood of capturing infection, particularly during early onset of an outbreak.

Our work is an important step towards primary care testing for COVID-19 infection. Firstly, our study is the first one to show that early RT-PCR testing in primary care can accurately detect COVID-19 infection. Secondly, the Austrian experience demonstrates that primary care can be a reliable partner for the control and prevention of COVID-19. Notably, countries with major epidemics, including the UK and the USA, have excluded primary care as an important testing site. Primary care in the UK has highly sophisticated linked searchable coded systems, meaning that prioritising testing in this setting rapidly creates exceptional real-time epidemiological data critical to identify risk groups and is capable of evaluating impacts of interventions such as social distancing and lockdown. Thirdly, evidence from countries like South Korea,^33^ where large-scale TTI strategies have been able to control the spread of COVID-19, and existing modelling studies in the UK^34^ highlight the need for comprehensive and effective TTI strategies to prevent onwards transmission of COVID-19 and suppress the virus preventing a secondary epidemic peak. Our study adds to this evidence and suggests that additional testing should be done in primary care and encouraged as early as symptoms appear. To our knowledge, our study is the first to highlight the need for large-scale and early primary care RT-PCR testing as part of an effective TTI strategy.

Our study has many strengths. Firstly, we included data from a well-established sentinel practice, participating in the National Influenza Screening Programme, covering a political subdistrict in Austria. Secondly, national SARS-CoV-2 screening was adopted early, starting the day before the first two cases were reported in Austria; and 16 of 29 cases documented in the Schladming-Dachstein region, including the first and the last case, were detected at the sentinel practice. The National health hotline 1450 and local general practitioners systematically referred people with mild to moderate symptoms to the sentinel practice, and patient self-referral was encouraged in the local media and on the practice home page. RT-PCR testing was rapidly deployed by offering same day GP appointments, and result reporting and case notification within 24 hours. To protect patients and staff, an internal safety protocol was developed to include physical separation of patients presenting with flu-like symptoms from those people receiving routine care, use of full personal protective equipment, and regular self-testing of staff. Rapid adoption of new commercial antibody platforms (Lab Mustafa, Salzburg) and in-house neutralising antibody testing assay (Medical University of Vienna) enabled accurate interpretation of RT-PCR results.

There are some limitations of our study. Firstly, we used a relatively small patient cohort from a single sentinel practice, potentially limiting conclusions on causality and generalisability of our finding to other areas and secondly, we excluded seven patients for whom COVID-19 serostatus were not available. Lack of association with high fever and cough in our COVID-19 cohort, may be due to the National health hotline 1450 directing patients with more severe disease to attend emergency service. Therefore, people with these symptoms might have preferred to attend acute services rather than general practice. Although we collected data prospectively, recall bias cannot be excluded. This could be suggested by the lack of association of symptoms of acute infection (nausea and vomiting, breathless and myalgia) among all people confirmed with infection (when including those with negative RT-PCR), compared to those people presenting early (reactive RT-PCR). Specific recall bias of taste is less likely, as it featured in both groups and data collection was completed prior to publication of the first systematic review of altered taste and smell in the media.^3^

To our knowledge, this is the first study to show that primary care can contribute to early case detection and termination of a SARS-CoV-2 outbreak in the community. Our study has important implications for patients, public health, and health systems; nationally and internationally for outbreak epidemiology and control. Strict adherence to safety protocols allows continuation of routine care, potentially reducing the chance of excess non-COVID-related and mortality among the practice population. As countries enter the viral suppression phase, early detection will be crucial in the prevention and control of the disease. Early testing at onset of disease, followed by timely contact tracing and case isolation of secondary cases should prevent onward transmission and reduce the reproduction number R below 1. Austria has increased the number of its sentinels sites from 91 to 231 due to COVID-19, indicating that primary care has become an essential partner in a comprehensive surveillance strategy for disease prevention and control. Key priorities for future research include generalisability of the intervention in multi-ethnic inner-city settings, systematic quantitative and qualitative evaluation of the Austrian National SARS-CoV-2 screening programme, and comparative analysis of the SARS-CoV-2 outbreak with the preceding seasonal influenza season.

## Data Availability

Data and codes for the modelling are available from the corresponding authors on request.

## Contribution

The WL, OL, MRF, MEMK, EMH, CH and JPG contributed to the design of the study. OL and EMH took nasopharyngeal swabs. OL, EMH and CH maintained the clinical data base. AS and RG submitted the ethics application. MRF provided RT-PCR data, MEMK produced ELISA data, and KS performed the neutralising antibody assay. JPG and WL conducted the statistical analysis. WL and JPG wrote the manuscript with contribution from OL, MRF, MEMK, RCG, EMH, CH, AS and CG. All authors read and approved the final version.

## Acknowledgments

We thank Evelyn Marktl for daily updates on the Christian Drosten’s COVID-19 podcast (https://www.ndr.de/nachrichten/info/podcast4684.html). We are grateful to the team of Praxis Dr Lammel for their contributions, and in particular to the nurse Sabine Roiderer for providing direct patient care and help with administration. We thank the patients of Schladming-Dachstein for participating in the study.

## References

1. World Health Organisation (WHO). Clinical management of severe acute respiratory infection when COVID-19 is suspected. 2020. https://www.who.int/publications-detail/clinical-management-of-severe-acute-respiratory-infection-when-novel-coronavirus-(ncov)-infection-is-suspected (accessed July 02, 2020)

2. Tong JY, Wong A, Zhu D, Fastenberg JH, Tham T. The Prevalence of Olfactory and Gustatory Dysfunction in COVID-19 Patients: A Systematic Review and Meta-analysis. Otolaryngol Head Neck Surg 2020: 194599820926473.

3. Lovato A, de Filippis C. Clinical Presentation of COVID-19: A Systematic Review Focusing on Upper Airway Symptoms. Ear Nose Throat J 2020: 145561320920762.

4. Lechner M, Chandrasekharan D, Jumani K, et al. Anosmia as a presenting symptom of SARS-CoV-2 infection in healthcare workers - A systematic review of the literature, case series, and recommendations for clinical assessment and management. Rhinology 2020.

5. Lauer SA, Grantz KH, Bi Q, et al. The Incubation Period of Coronavirus Disease 2019 (COVID-19) From Publicly Reported Confirmed Cases: Estimation and Application. Ann Intern Med 2020; 172(9): 577–82.

6. Cheng HY, Jian SW, Liu DP, Ng TC, Huang WT, Lin HH. Contact Tracing Assessment of COVID-19 Transmission Dynamics in Taiwan and Risk at Different Exposure Periods Before and After Symptom Onset. JAMA Intern Med 2020.

7. Kimball A, Hatfield KM, Arons M, et al. Asymptomatic and Presymptomatic SARS-CoV-2 Infections in Residents of a Long-Term Care Skilled Nursing Facility - King County, Washington, March 2020. MMWR Morb Mortal Wkly Rep 2020; 69(13): 377–81.

8. Bullard J, Dust K, Funk D, et al. Predicting infectious SARS-CoV-2 from diagnostic samples. Clin Infect Dis 2020.

9. Wölfel R, Corman VM, Guggemos W, et al. Virological assessment of hospitalized patients with COVID-2019. Nature 2020; 581(7809): 465–9.

10. Kay J. COVID-19 Superspreader Events in 28 Countries: Critical Patterns and Lessons. Quillette. 2020. https://quillette.com/2020/04/23/covid-19-superspreader-events-in-28-countries-critical-patterns-and-lessons/ (accessed July 02, 2020)

11. Endo A, Abott S, Aj K, Funk S. Estimating the overdispersion in COVID-19 transmission using outbreak sizes outside China. 2020. https://d212y8ha88k086.cloudfront.net/manuscripts/17377/bbd75255-2e3a-4f30-96c9-28142e604f0f_15842_-_akira_endo.pdf?doi=10.12688/wellcomeopenres.15842.1&numberOfBrowsableCollections=5&numberOfBrowsableInstitutionalCollections=0&numberOfBrowsableGateways=13 (accessed July 02, 2020)

12. Liu Y, Yan LM, Wan L, et al. Viral dynamics in mild and severe cases of COVID-19. Lancet Infect Dis 2020.

13. European Centre for Disease Control and Prevention (ECDC). Rapid Risk Assessment: Coronavirus disease 2019 (COVID-19) in the EU/EEA and the UK– ninth update. 2020. https://www.ecdc.europa.eu/en/publications-data/rapid-risk-assessment-coronavirus-disease-2019-covid-19-pandemic-ninth-update (accessed July 02, 2020)

14. Koo JR, Cook AR, Park M, et al. Interventions to mitigate early spread of SARS-CoV-2 in Singapore: a modelling study. Lancet Infect Dis 2020.

15. Frieden TR, Lee CT. Identifying and Interrupting Superspreading Events-Implications for Control of Severe Acute Respiratory Syndrome Coronavirus 2. Emerg Infect Dis 2020; 26(6): 1059–66.

16. Felbermayr G, Hinz J, Chowdhry S. Après-ski: The Spread of Coronavirus from Ischgl through Germany. CEPR Press - COVID ECONOMICS 2020 2020; 22.

17. Independent T 2020. https://www.independent.co.uk/news/world/europe/coronavirus-austria-cases-covid-19-hospital-lockdown-latest-a9466281.html. (accessed July 02, 2020)

18. (European Centre for Disease Control and Prevention (ECDC). Coronavirus disease 2019 (COVID-19) in the EU/EEA and the UK –ninth update, 2020. https://www.ecdc.europa.eu/sites/default/files/documents/covid-19-rapid-risk-assessment-coronavirus-disease-2019-ninth-update-23-april-2020.pdf (Accessed July 02, 2020)

19. de Sutter A, Llor C, Maier M, et al. Family medicine in times of ‘COVID-19’: A generalists’ voice. Eur J Gen Pract 2020; 26(1): 58–60.

20. Hull SA, Williams C, Ashworth M, Carvalho C, Boomla K. Suspected COVID-19 in primary care: how GP records contribute to understanding differences in prevalence by ethnicity. medRxiv 2020: 2020.05.23.20101741.

21. Marshall M, Howe A, Howsam G, Mulholland M, Leach J. COVID-19: a danger and an opportunity for the future of general practice. Br J Gen Pract 2020.

22. European Centres for Disease Control (ECDC). Strategies for the surveillance of COVID-19, 2020. https://www.ecdc.europa.eu/sites/default/files/documents/COVID-19-surveillance-strategy-9-Apr-2020.pdf (accessed July 11, 2020)

23. de Lusignan S, Dorward J, Correa A, et al. Risk factors for SARS-CoV-2 among patients in the Oxford Royal College of General Practitioners Research and Surveillance Centre primary care network: a cross-sectional study. Lancet Infect Dis 2020.

24. Zentrum für Virologie Medizinische Universität Wien. Projekt Diagnostisches Influenzanetzwerk Österreich (DINÖ). https://www.virologie.meduniwien.ac.at/wissenschaft-forschung/virus-epidemiologie/influenza-projekt-diagnostisches-influenzanetzwerk-oesterreich-dinoe/ (Accessed July 02, 2020)

25. Federal Ministry of Social Affairs H, Care and Consumer Protection, Republic of Austria. National Health Hotline 1450. 2019. https://www.1450.at/1450-die-gesundheitsnummer/ (accessed May 28, 2020).

26. Watson J, Whiting PF, Brush JE. Interpreting a covid-19 test result. BMJ 2020; 369: m1808.

27. Corman V, Bleicker T, Bru□nink S, et al. Diagnostic detection of 2019-nCoV by real-time RT-PCR, 2020. https://www.who.int/docs/default-source/coronaviruse/protocol-v2-1.pdf?sfvrsn=a9ef618c_2 (accessed July 02, 2020)

28. Stadlbauer D, Amanat F, Chromikova V, et al. SARS-CoV-2 Seroconversion in Humans: A Detailed Protocol for a Serological Assay, Antigen Production, and Test Setup. Curr Protoc Microbiol 2020; 57(1): e100.

29. Ahn JY, Sohn Y, Lee SH, et al. Use of Convalescent Plasma Therapy in Two COVID-19 Patients with Acute Respiratory Distress Syndrome in Korea. J Korean Med Sci 2020; 35(14): e149.

30. Pinnock H, Epiphaniou E, Sheikh A, et al. Developing standards for reporting implementation studies of complex interventions (StaRI): a systematic review and e-Delphi. Implementation Science 2015; 10(1): 42.

31. Menni C, Valdes AM, Freidin MB, et al. Real-time tracking of self-reported symptoms to predict potential COVID-19. Nat Med 2020.

32. He X, Lau EHY, Wu P, et al. Temporal dynamics in viral shedding and transmissibility of COVID-19. Nat Med 2020; 26(5): 672–5.

33. Shim E, Tariq A, Choi W, Lee Y, Chowell G. Transmission potential and severity of COVID-19 in South Korea. Int J Infect Dis 2020; 93: 339–44.

34. Panovska-Griffiths, Kerr CC, Stuard RM, Mistry D, Klein DJ, Viner RM, Bonnell CM. Determining the optimal strategy for reopening schools, workplaces and society in the UK: modelling patterns of reopening, the impact of test and trace strategies and risk of occurrence of a secondary COVID-19 pandemic wave. Lancet Child & Adolescent Health (in press); preprint on medRxiv doi: 10.1101/2020.06.01.20100461

